# A Cross-Cohort Validated Plasma Lipid Biomarker Assay for Early Breast Cancer Detection Using Machine Learning

**DOI:** 10.64898/2026.04.23.26351564

**Authors:** Tim Huang, Forrest C. Koch, David A. Peake, Klaus-Peter Adam, Mark David, Desmond Li, Kerry Heffernan, Ameline Lim, John G. H. Hurrell, Simon Preston, Amani Baterseh, Fatemeh Vafaee

## Abstract

Early detection of breast cancer remains essential for improving clinical outcomes, and complementary non-invasive approaches are needed to support existing screening methods, particularly for women with dense breast tissue. We have previously reported plasma lipid biomarker discovery using untargeted high-resolution liquid chromatography tandem mass spectrometry (LC–MS/MS). In this study, we performed biomarker confirmation and developed machine-learning models applied to targeted plasma lipid measurements for the non-invasive detection of early-stage breast cancer across international cohorts with independent external validation. Targeted LC–MS/MS was used to quantify candidate lipid panels in plasma samples from European discovery cohorts (n = 554) and an independent Australian cohort (n = 266) used for external validation. Data-driven feature selection identified a 15-lipid panel with strong performance in European cohorts (AUC ≥ 0.94). External validation prior to confidence stratification yielded 76% sensitivity, 64% specificity, and an AUC of 0.81 in the Australian validation cohort. Clinical assay development requires iterative panel and model testing to support translational feasibility and performance in the intended-use population. An analytically viable panel, excluding lipids requiring complex and costly synthesis, achieved comparable accuracy with improved assay robustness. Confidence-based analysis showed enhanced performance for predictions made with moderate to high confidence, with sensitivity up to 89% and AUC up to 0.85, suggesting that ongoing research should focus on strategies to enhance diagnostic model confidence. Importantly, model predictions were independent of breast density, tumour size, grade, subtype, and morphology, indicating biological specificity of the lipid signature. These results demonstrate that calibrated machine-learning models applied to plasma lipid biomarkers can support non-invasive breast cancer detection. Expanding training datasets to include greater diversity will further improve performance in the ongoing development of this lipid-based detection approach.

## Introduction

Breast cancer remains a major global health challenge, with more than 2.26 million new cases and 685,000 deaths reported in 2020, a burden projected to rise to 3 million cases and 1 million deaths annually by 2040^1,2^. Despite advances in precision oncology and targeted therapeutics, mortality continues to increase across the globe, in part reflecting the challenges of the disease’s clinical and molecular heterogeneity^3,4^. Access to advanced diagnostic and treatment modalities is also uneven, with disadvantaged populations experiencing disproportionately poorer outcomes^5,6^. Consequently, early detection remains one of the most effective strategies for reducing breast cancer mortality.

Current screening relies largely on mammography and ultrasound, yet these modalities have reduced sensitivity in women with dense breast tissue^7,8^, and have a false-positive rate that frequently lead to unnecessary biopsies^9^. Although biopsy remains the diagnostic gold standard, it is invasive, resource-intensive, and unsuitable for widespread population-level screening. These limitations have motivated interest in blood-based “liquid biopsies” that offer minimally-invasive, scalable alternatives capable of capturing disease-associated molecular signatures^10^.

Among circulating biomarkers, lipids have emerged as promising candidates for cancer detection^11,12^ due to their roles in membrane remodelling, energy metabolism, and oncogenic signalling. Furthermore, lipid profiles in the breast tumour microenvironment are distinctly different to that in normal breast tissue^13^ providing evidence to support a biological link between blood-lipid profiles and the presence of breast cancer in women. Advances in liquid chromatography coupled with high-resolution and tandem mass spectrometry (LC–MS/MS) now enable sensitive and reproducible measurement of plasma-derived lipid species at scale. Indeed, we have previously published a plasma lipid biomarker discovery study using this technology to differentiate between women with and without breast cancer^12^. Candidate biomarkers selected using untargeted, high-resolution LC–MS/MS must be confirmed using targeted LC–MS/MS.

In this study, we used a candidate lipid panel identified using untargeted, high-resolution LC–MS/MS in previous work from our group^12^, which was subsequently refined to meet both identification and method qualification criteria^14^. Interpreting high-dimensional lipidomic profiles requires computational approaches capable of detecting subtle multivariate patterns not discernible through manual inspection. Machine learning (ML) provides a data-driven framework to identify lipid combinations that optimally discriminate cancer from control samples. Using a nonlinear multivariate ML strategy, features with consistent discriminative value were identified across European discovery cohorts, resulting in a 15-lipid panel with strong performance in held-out European cohorts. Model generalisability was then assessed using an independent Australian cohort collected in a distinct clinical and demographic setting to evaluate cross-population applicability. The panel was subsequently refined using complementary considerations, including reduction of feature collinearity and analytical viability, to improve assay suitability while maintaining predictive performance. Together, these findings provide a systematic framework for translating lipid biomarkers into clinically viable diagnostic assays.

## Results

### Cohort Characteristics and Sample Composition

Fasted peripheral blood samples were prospectively collected from women who were assigned female at birth. Plasma from these blood samples was used for all analyses in this study. These participants were recruited between 2020 and 2023 across multiple clinical sites in Eastern Europe and Australia (n=820), which included 406 women with recently diagnosed breast cancer and 414 control women with no current or previous breast cancer diagnosis. All breast cancer diagnoses were confirmed by tissue biopsy. Four independent cohorts were included in this study (**Table 1**). Three European cohorts referred to as EU-cohort 1 (n = 153), EU-cohort 2 (n = 199), and EU-cohort 3 (n = 202), were collected sequentially and together represent a temporally diverse discovery dataset. Donor demographics and disease characteristics were recorded, including age, body mass index, smoking status, breast cancer type, and stage (**Tables 2-4**). Breast cancer morphology and molecular subtypes were recorded where data was available.

**Table 1.**
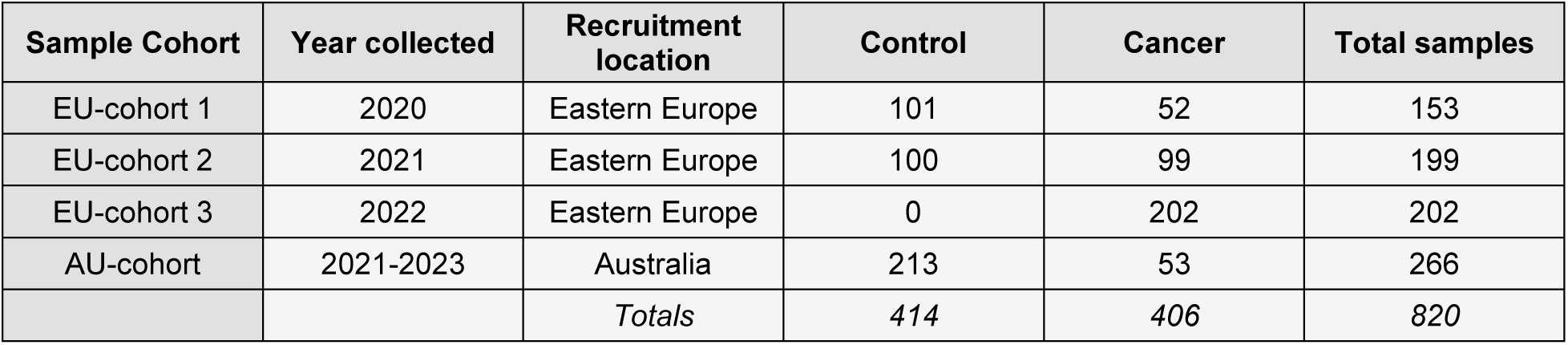
The datasets that were used in this study, along with their breakdown by cohort. The year of sample collection, subject recruitment location and number of samples broken down by tumour status is indicated.

**Table 2.** Comparison of key clinical features between cancer and controls in each cohort. The statistical tests are based on the chi-squared test of independence and an ANOVA for categorical and continuous data, respectively.NA, not applicable; NR, not recorded.

| Demographics and disease characteristics | Cohort |  | Disease state |  | P-value |
| --- | --- | --- | --- | --- | --- |
|  |  |  | Cancer | Control |  |
| Age | EU-cohort 1 | 25<sup>th</sup> % | 51 | 48 | 1.4e<sup>-03</sup> |
|  |  | Median | 58 | 52 |  |
|  |  | 75<sup>th</sup> % | 68 | 59 |  |
|  | EU-cohort 2 | 25<sup>th</sup> % | 49 | 47 | 0.58 |
|  |  | Median | 57 | 56 |  |
|  |  | 75<sup>th</sup> % | 65 | 64 |  |
|  | EU-cohort 3 | 25<sup>th</sup> % | 52 | NA | NA |
|  |  | Median | 62 | NA |  |
|  |  | 75<sup>th</sup> % | 69 | NA |  |
|  | AU-cohort | 25<sup>th</sup> % | 51 | 48 | 4.1e<sup>-03</sup> |
|  |  | Median | 59 | 53 |  |
|  |  | 75<sup>th</sup> % | 70 | 61 |  |
| BMI | EU-cohort 1 | 25<sup>th</sup> % | 24.9 | 23.8 | 3.0e<sup>-02</sup> |
|  |  | Median | 27.6 | 25.6 |  |
|  |  | 75<sup>th</sup> % | 32.3 | 29.4 |  |
|  | EU-cohort 2 | 25<sup>th</sup> % | 24.1 | 24.3 | 0.71 |
|  |  | Median | 27.2 | 27.4 |  |
|  |  | 75<sup>th</sup> % | 29.7 | 29.9 |  |
|  | EU-cohort 3 | 25<sup>th</sup> % | 24.2 | NA | NA |
|  |  | Median | 27.6 | NA |  |
|  |  | 75<sup>th</sup> % | 30.8 | NA |  |
|  | AU- cohort | 25<sup>th</sup> % | 24.4 | 22.2 | 3.9e<sup>-6</sup> |
|  |  | Median | 27.5 | 24.8 |  |
|  |  | 75<sup>th</sup> % | 34.7 | 28.1 |  |
| Smoking history | EU-cohort 1 | Yes | 4 | 15 | 0.31 |
|  |  | No | 48 | 86 |  |
|  | EU-cohort 2 | Yes | 1 | 10 | 1.4e<sup>-02</sup> |
|  |  | No | 98 | 90 |  |
|  |  | Yes | 6 | NA |  |
|  | **EU-cohort 3** | **No** | 196 | NA | NA |
|  | **AU-cohort** | **Yes** | NR | NR | NA |
|  |  | **No** | NR | NR | NA |

**Table 3.**
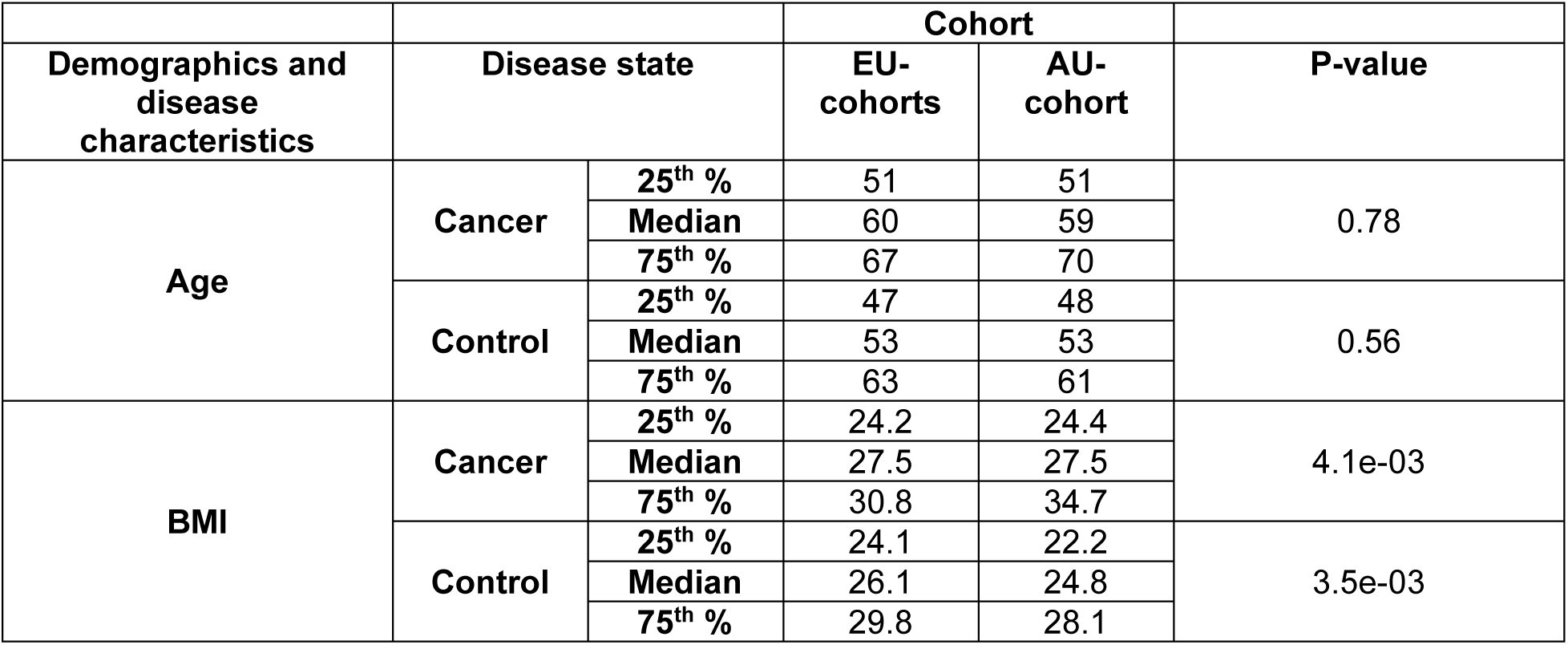
Comparison of key clinical features between cancer and controls across pooled EU and AU cohorts. The statistical tests are based on an ANOVA.

| Demographics and disease characteristics | Disease state |  | Cohort |  | P-value |
| --- | --- | --- | --- | --- | --- |
|  |  |  | EU-cohorts | AU-cohort |  |
| Age | Cancer | **25<sup>th</sup> %** | 51 | 51 | 0.78 |
|  |  | **Median** | 60 | 59 |  |
|  |  | **75<sup>th</sup> %** | 67 | 70 |  |
|  | Control | **25<sup>th</sup> %** | 47 | 48 | 0.56 |
|  |  | **Median** | 53 | 53 |  |
|  |  | **75<sup>th</sup> %** | 63 | 61 |  |
| BMI | Cancer | **25<sup>th</sup> %** | 24.2 | 24.4 | 4.1e-03 |
|  |  | **Median** | 27.5 | 27.5 |  |
|  |  | **75<sup>th</sup> %** | 30.8 | 34.7 |  |
|  | Control | **25<sup>th</sup> %** | 24.1 | 22.2 | 3.5e-03 |
|  |  | **Median** | 26.1 | 24.8 |  |
|  |  | **75<sup>th</sup> %** | 29.8 | 28.1 |  |

**Table 4.**
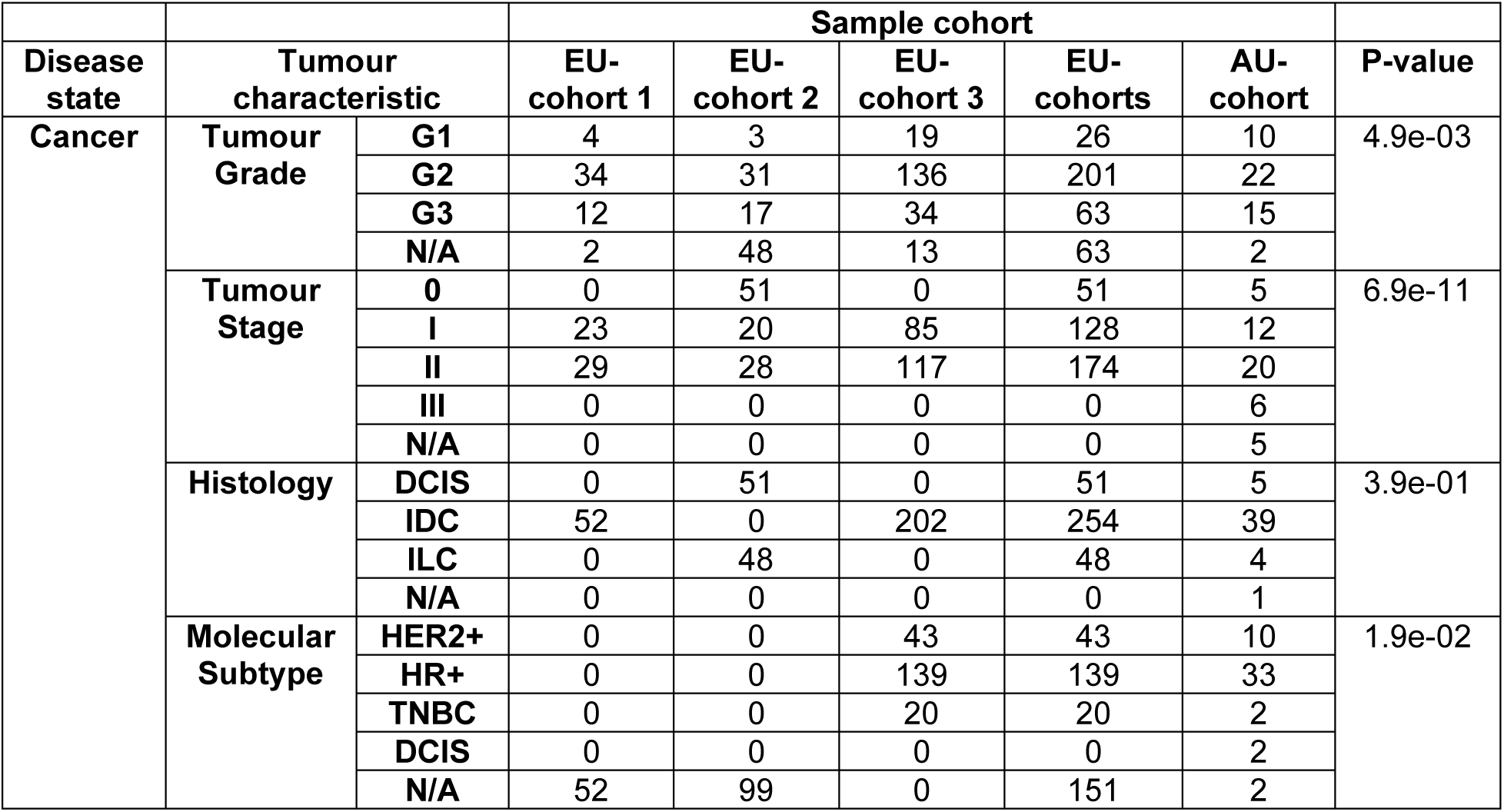
Comparison of key clinical features across cohorts. The statistical tests are based on the chi-squared test of independence comparing the combined EU-cohort to the AU-cohort.

| Disease state | Tumour characteristic |  | Sample cohort |  |  |  |  | P-value |
| --- | --- | --- | --- | --- | --- | --- | --- | --- |
|  |  |  | EU-cohort 1 | EU-cohort 2 | EU-cohort 3 | EU-cohorts | AU-cohort |  |
| Cancer | Tumour Grade | G1 | 4 | 3 | 19 | 26 | 10 | 4.9e-03 |
|  |  | G2 | 34 | 31 | 136 | 201 | 22 |  |
|  |  | G3 | 12 | 17 | 34 | 63 | 15 |  |
|  |  | N/A | 2 | 48 | 13 | 63 | 2 |  |
|  | Tumour Stage | 0 | 0 | 51 | 0 | 51 | 5 | 6.9e-11 |
|  |  | I | 23 | 20 | 85 | 128 | 12 |  |
|  |  | II | 29 | 28 | 117 | 174 | 20 |  |
|  |  | III | 0 | 0 | 0 | 0 | 6 |  |
|  |  | N/A | 0 | 0 | 0 | 0 | 5 |  |
|  | Histology | DCIS | 0 | 51 | 0 | 51 | 5 | 3.9e-01 |
|  |  | IDC | 52 | 0 | 202 | 254 | 39 |  |
|  |  | ILC | 0 | 48 | 0 | 48 | 4 |  |
|  |  | N/A | 0 | 0 | 0 | 0 | 1 |  |
|  | Molecular Subtype | HER2+ | 0 | 0 | 43 | 43 | 10 | 1.9e-02 |
|  |  | HR+ | 0 | 0 | 139 | 139 | 33 |  |
|  |  | TNBC | 0 | 0 | 20 | 20 | 2 |  |
|  |  | DCIS | 0 | 0 | 0 | 0 | 2 |  |
|  |  | N/A | 52 | 99 | 0 | 151 | 2 |  |

To evaluate the candidate lipid panels, models and the potential of a lipid-based breast cancer detection assay, the generalizability across populations and collection environments, were evaluated using an external dataset/cohort (AU-cohort) comprising 266 samples. These samples were obtained from prospectively recruited women from clinical sites in Australia. This cohort is geographically and temporally distinct from the EU-cohorts and was used exclusively as an external validation dataset.

Together, the four cohorts comprise 414 controls and 406 early-stage cancers. The balanced disease representation and multi-regional sampling framework are critical to evaluating the stability, reproducibility, and portability of lipid-based liquid biopsy signature across diverse populations.

### Biomarker Panel Identification and Breast Cancer Detection Pipeline

The purpose of this study was to provide biomarker confirmation and subsequent development of a plasma lipid-based assay, coupled with machine-learning models for the non-invasive detection of early-stage breast cancer. The European cohorts had previously been used for feature selection and the evaluation of potential diagnostic value in breast cancer using untargeted LC-MS/MS^12^. Although the discovery of potential lipid biomarkers is an important step toward the development of a clinical assay, the ability to accurately and reproducibly quantify these lipids is essential to ongoing development. To this end, we have also performed lipid identification and developed targeted LC-MS/MS method to measure the concentrations of candidate biomarkers in plasma to enable the accurate quantification of lipids in this study^14^.

This study applied a machine-learning–driven strategy to identify a plasma lipid biomarker panel capable of distinguishing early-stage breast cancer from healthy controls. The overall workflow (**Figure 1A**) integrates data preprocessing, data-driven biomarker discovery, and ensemble machine-learning modelling, followed by multi-cohort validation across European and Australian populations.

**Figure 1.**
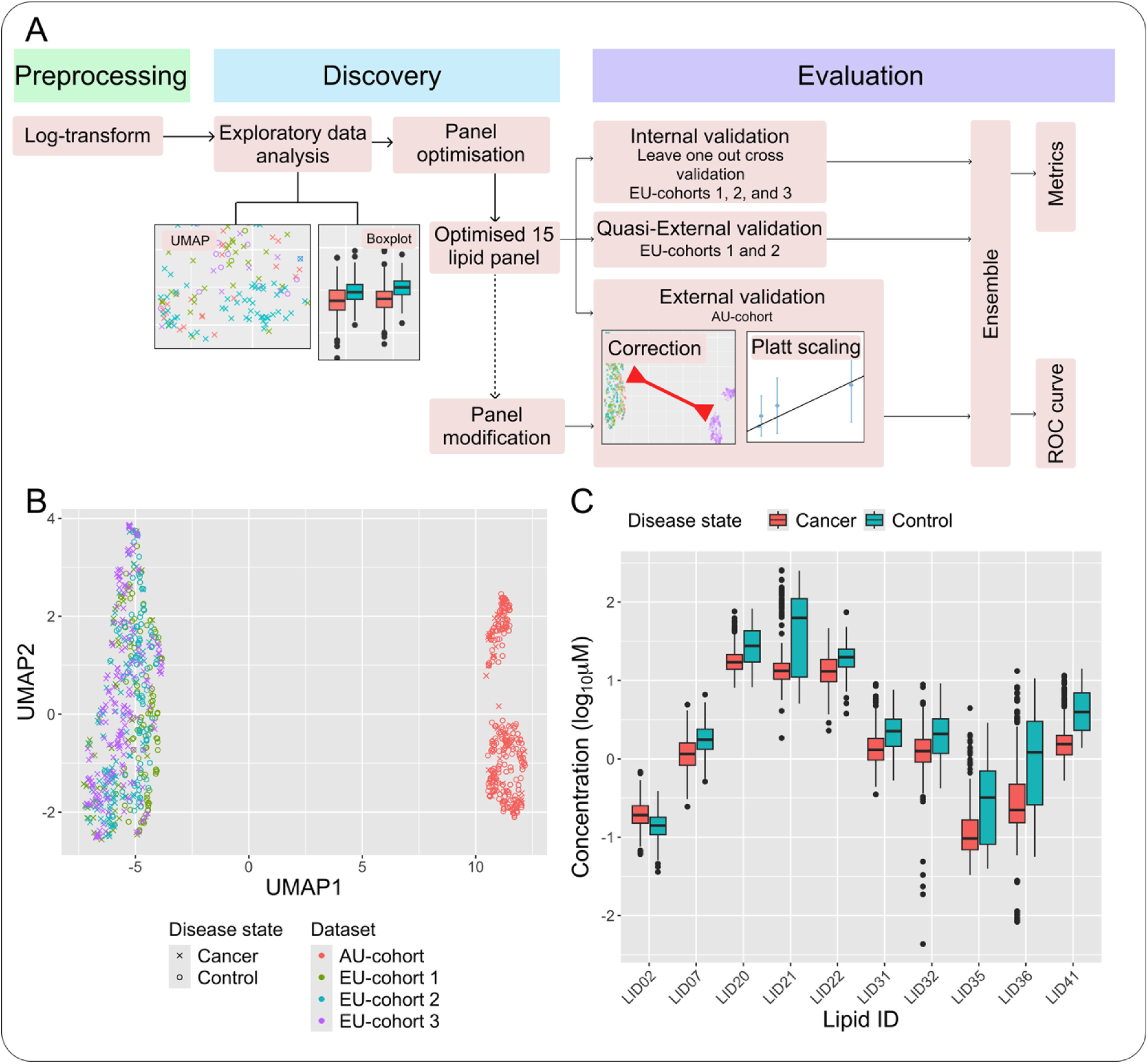
(A) The discovery and evaluation pipeline. (B) An initial EDA is presented in the UMAP, and (C) a boxplot representation of the top 10 lipids with significant differences between disease states. EDA, Exploratory data analysis; UMAP, Uniform Manifold Approximation and Projection plot.

All lipid concentrations were log-transformed to stabilise variance and reduce the influence of outliers. Biomarker discovery was performed on the European cohorts (EU-cohort 1-3) using a fully data-driven feature-selection framework. This approach evaluates one thousand bootstrapped subsets of data to identify lipids that consistently differ between cancer and control groups, thereby yielding a robust candidate biomarker panel that is less sensitive to sampling variability.

The resulting lipid panel was then evaluated within a structured validation framework. Internal validation was performed using leave-one-out cross-validation (LOOCV) in EU-cohorts 1 – 3, while quasi-external validation assessed performance on held-out European datasets. Crucially, clinical generalisability was tested using an external Australian cohort (AU-cohort), which was geographically and temporally distinct from the discovery cohorts. Model performance was assessed using sensitivity, specificity, accuracy, F1-score, and AUC to provide a comprehensive view of diagnostic utility.

To generate these diagnostic estimates, we trained an optimised ensemble machine-learning model, which combines predictions from several complementary algorithms. Ensemble models generally outperform individual models because they capture different aspects of the data, offer greater stability, and mitigate model-specific biases. Here, the ensemble was selected and refined using a smaller development dataset to retain only models that contributed meaningful and non-redundant predictive information.

Initial exploration of the data showed that the discovery dataset (EU-cohort 1-3) exhibited no major batch-related differences, and cancer and control samples displayed a degree of nonlinear separation that could be effectively captured through our biomarker discovery and machine-learning modelling framework. In contrast, the AU-cohort formed a clearly distinct cluster (**Figure 1B**), indicating a substantial batch difference, likely driven by geographic, temporal, and technical factors. This separation suggested that batch-correction procedures may be appropriate.

### Data-Driven Optimisation of a Plasma Lipid Biomarker Panel

This study attempted to measure the concentrations of 48 potential lipid biomarkers in plasma across all discovery cohorts (**Table 5**). Many of these analytes showed little or no variation between cancer and control groups, and their inclusion could introduce noise into machine-learning models, reduce interpretability, and increase analytical burden. Refining the panel was therefore essential both for improving model performance and for reducing downstream costs associated with assay development and clinical translation.

**Table 5.** Lipid analytes used in this study. LID, lipid identifier; Y, yes; N, no.

| LID | Lipid Annotation | Qualified (Y/N) |
| --- | --- | --- |
| 01 | CAR 18:2 | Y |
| 02 | CER d18:1/18:0 | Y |
| 03 | CER d18:1/24:0 | Y |
| 04 | LPA 16:0 | N |
| 05 | LPA 18:0 | N |
| 06 | LPA 18:2 | Y |
| 07 | LPC 14:0 | Y |
| 08 | LPC 16:0 | Y |
| 09 | LPC 18:0 | Y |
| 10 | LPC 18:2 | Y |
| 11 | LPC O-16:0 | Y |
| 12 | LPE 18:0 | Y |
| 13 | LPE 22:6 | Y |
| 14 | LPI 18:1 | N |
| 15 | PC 14:0/16:0 | Y |
| 16 | PC 16:0/16:1 | Y |
| 17 | PC 16:1/16:1 | N |
| 18 | PC 18:0/18:0 | Y |
| 19 | PC 18:0/18:2 | Y |
| 20 | PC 18:1/18:1 | Y |
| 21 | PC 18:1/20:4 | Y |
| 22 | PC 18:2/18:2 | Y |
| 23 | PE 16:0/18:1 | Y |
| 24 | PE 16:0/20:4 | Y |
| 25 | PE 18:0/18:2 | Y |
| 26 | PE O-18:0/20:4 | Y |
| 27 | PE P-16:0/18:2 | Y |
| 28 | PE P-18:0/20:4 | Y |
| 29 | PE P-18:0/22:6 | Y |
| 30 | PG 18:0/18:1 | N |
| 31 | PI 16:0/18:1 | Y |
| 32 | PI 18:0/18:1 | Y |
| 33 | PI 18:0/20:4 | Y |
| 34 | PI 18:2/20:4 | N |
| 35 | PS 18:0/18:1 | N |
| 36 | PS 18:0/20:4 | N |
| 37 | SM d18:1/17:0 | N |
| 38 | SM d18:1/18:0 | Y |
| 39 | SM d18:1/18:1 | Y |
| 40 | SM d18:1/24:0 | Y |
| 41 | SPHP d18:1 | Y |
| 42 | TG 16:0/18:1/23:0 | Y |
| 43 | TG 18:1/18:1/18:2 | Y |
| 44 | TG 18:1/18:1/22:0 | Y |
| 45 | TG 18:1/18:1/22:1 | Y |
| 46 | TG 18:2/18:2/18:1 | Y |
| 47 | TG 18:2/18:2/18:2 | Y |
| 48 | TG O-16/18:1/18:2 | Y |

To identify the most informative biomarkers, we employed a data-driven feature-selection strategy. We applied the Boruta^15^ algorithm (i.e., a wrapper method built around a random forest classifier) to evaluate lipid biomarker importance relative to synthetic “shadow” features representing random noise. By repeating Boruta over 1,000 bootstrap resamples of the discovery dataset (EU-cohort 1-3), we selected lipids that remained informative across population subsets while minimising the influence of sampling variability and outliers. The log-transformed plasma-lipid measurements of the top 10 most differentially expressed lipids between groups illustrates the concentration range of lipids that together may be able to discriminate between breast cancer and control plasma samples (**Figure 1C**).

Lipids selected in more than 70% of bootstrap iterations were considered robust candidates. Twenty candidate lipids met this criterion (**Figure 2A**). Five lipids were subsequently excluded, as their identities could not be confidently confirmed in plasma and/or they did not meet our method qualification criteria defined in our former study^14^ (i.e., SM d18:1/17:0, LPI 18:1, LPA 18:0, LPA 18:2 and PS 18:0/20:4), resulting in an optimised panel of 15 lipids (**Table 6**).

**Figure 2.**
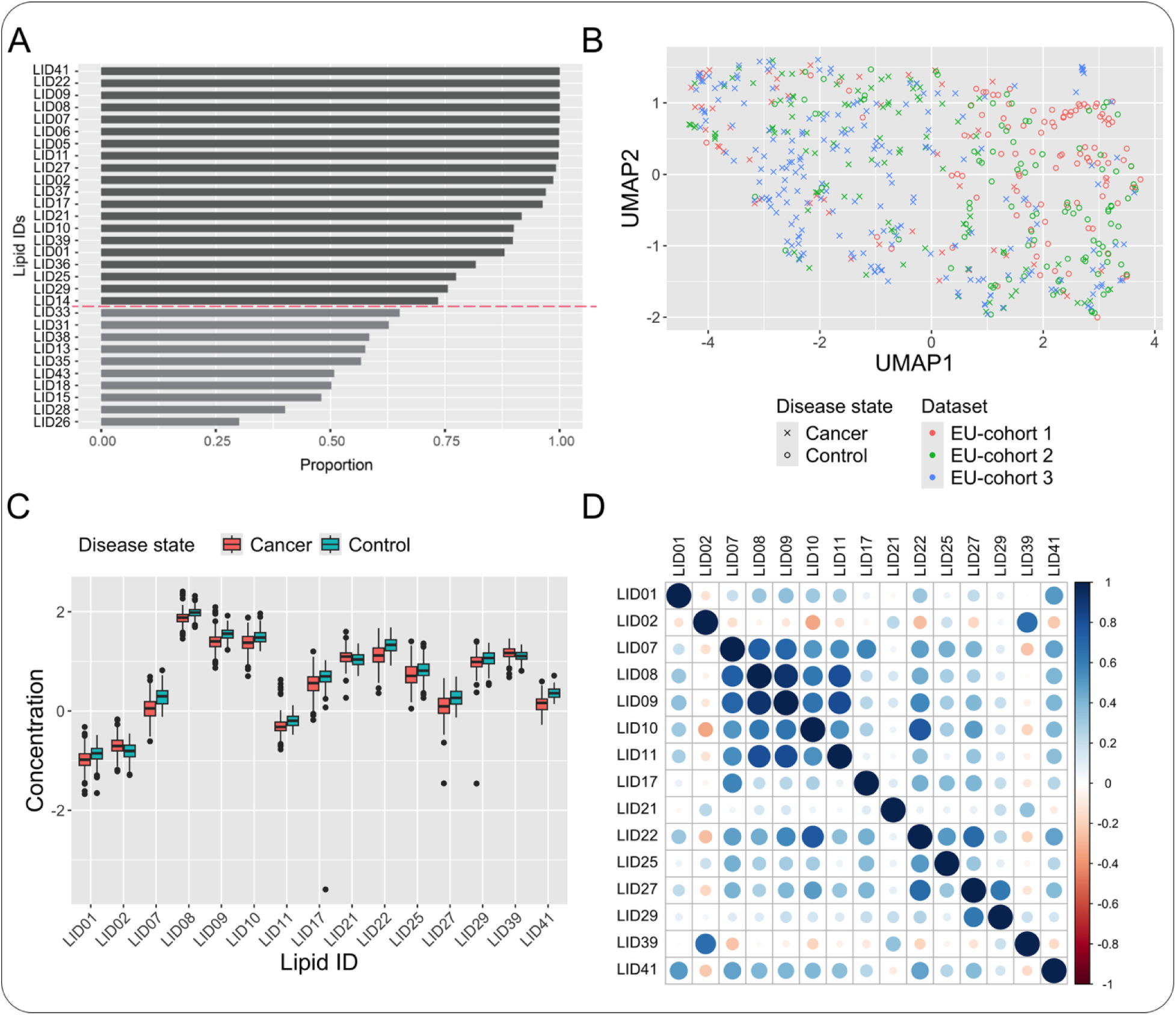
(A) Boruta analysis ranked the top 30 lipids in the order of importance to predict breast cancer vs control. Lipids that were identified as important in at least 70% of the bootstraps were selected (20 lipids). The red dotted line separates the top 20. (B) The UMAP of EU-cohort 1-3 using the optimised panel (15 lipids). (C) Boxplot graph of log transformed lipid concentrations in the optimised panel stratified by disease state. (D) Correlation matrix indicating degrees of collinearity between lipids from the optimised panel.

**Table 6.** Composition of lipid panels. Y, yes (in panel), N, no (not in panel).

| LID | Lipid name | Lipid panel name |  |  |  |
| --- | --- | --- | --- | --- | --- |
|  |  | Optimised | Minimal viable | Low collinearity | Analytically viable |
| 01 | CAR 18:2 | Y | N | Y | Y |
| 02 | CER d18:1/18:0 | Y | N | Y | Y |
| 07 | LPC 14:0 | Y | Y | N | Y |
| 08 | LPC 16:0 | Y | Y | N | Y |
| 09 | LPC 18:0 | Y | Y | Y | Y |
| 10 | LPC 18:2 | Y | N | Y | Y |
| 11 | LPC O-16:0 | Y | N | Y | Y |
| 17 | PC 16:1/16:1 | Y | N | Y | Y |
| 21 | PC 18:1/20:4 | Y | Y | Y | N |
| 22 | PC 18:2/18:2 | Y | Y | Y | Y |
| 25 | PE 18:0/18:2 | Y | N | Y | Y |
| 27 | PE P-16:0/18:2 | Y | N | Y | Y |
| 29 | PE P-18:0/22:6 | Y | N | Y | Y |
| 39 | SM d18:1/18:1 | Y | N | Y | Y |
| 41 | SPHP d18:1 | Y | Y | Y | Y |

This optimised, data-driven panel formed the foundation for downstream model training, validation, and panel-modification strategies. Exploratory visualisation of these 15 lipids demonstrated separation between cancer and control samples in the discovery cohorts. UMAP embeddings showed an improved clustering of disease states (**Figure 2B**), and the boxplots comparing lipid concentrations in cancers and controls reflect this (**Figure 2C**). Although the overall collinearity in the optimised panel is small, some lipids were highly correlated (**Figure 2D**).

### Ensemble Model Construction and Refinement

The predictive model was implemented as an ensemble comprising 11 complementary machine-learning algorithms (model specifications, R functions, and hyperparameters are provided in **Table 7**). Each algorithm generated a prediction score between 0 (control) and 1 (cancer), and the ensemble output was calculated as the mean of all individual model predictions. Ensemble averaging generally improves robustness by leveraging diverse modelling assumptions and reducing the influence of any single poorly performing algorithm.

**Table 7.** The machine learning models in the optimised ensemble. Their R functions and optimized hyperparameters are also provided. Models with “NA” entries have been removed from the optimized ensemble. NA, not applicable.

| Model | R function |  | Hyperparameters by panel |  |  |  |
| --- | --- | --- | --- | --- | --- | --- |
|  |  |  | Mix0 | Minimal viable | Non-collinear | Analytically viable |
| Support Vector Machine with Radial Basis Function Kernel | svmRadial | sigma | 9.82e-03 | 1.08e-01 | 1.65e-01 | 3.20e-01 |
|  |  | C | 6.19e+00 | 1.25e+01 | 4.81e-01 | 1.17e-01 |
|  | svmRadialSigma | sigma | 3.37e-02 | 3.43e-02 | 3.45e-02 | 1.95e-02 |
|  |  | C | 1.96e+01 | 1.61e+02 | 1.93e+00 | 4.56e+00 |
| Model Averaged Neural Network | avNNet | size | 1.00e+01 | 4.00e+00 | 1.00e+01 | 1.00e+01 |
|  |  | decay | 6.43e-05 | 8.75e-02 | 6.43e-05 | 6.43e-05 |
|  |  | bag | True | False | True | True |
| K-Nearest Neighbors | knn | k | 1.23e+02 | 3.50e+01 | 1.21e+02 | 2.10e+01 |
| Adjacent Categories Probability Model for Ordinal Data | vglmAdjCat | parallel | True | False | False | False |
|  |  | link | loge | loge | loge | loge |
| Cumulative Probability Model for Ordinal Data | vglmCumulative | parallel | True | True | False | False |
|  |  | link | logit | cauchit | probit | probit |
| eXtreme Gradient Boosting | xgbTree | nrounds | 5.11e+02 | 9.50e+02 | 5.52e+02 | 4.30e+02 |
|  |  | max_depth | 9.00e+00 | 9.00e+00 | 4.00e+00 | 9.00e+00 |
|  |  | eta | 4.45e-01 | 3.32e-01 | 4.87e-01 | 3.93e-01 |
|  |  | gamma | 1.47e+00 | 5.72e+00 | 5.72e-03 | 6.62e+00 |
|  |  | colsample_bytree | 6.42e-01 | 4.04e-01 | 5.98e-01 | 6.23e-01 |
|  |  | min_child_weight | 1.30e+01 | 1.00e+01 | 1.20e+01 | 1.00e+00 |
|  |  | subsample | 9.56e-01 | 5.98e-01 | 8.55e-01 | 7.46e-01 |
| Stochastic Gradient Boosting | gbm | NA | NA | NA | NA | NA |
| Multi-Layer Perceptron | mlp | NA | NA | NA | NA | NA |
| Distance Weighted Discrimination with Polynomial Kernel | dwdPoly | NA | NA | NA | NA | NA |
| Distance Weighted Discrimination with Radial Basis Function Kernel | dwdRadial | NA | NA | NA | NA | NA |

Although the constituent models were initially selected based on their complementary predictive mechanisms and suitability for high-dimensional biological data, empirical behaviour during training showed that some algorithms generated unstable or uninformative predictions. To maintain a reliable ensemble, we systematically assessed each model’s prediction distribution and removed those that could adversely affect downstream performance.

The ensemble was trained on EU-cohort 1–3 with the optimised panel, and training predictions were generated per ML model. The boxplots of the training predictions are shown in **Figure 3A**, visualising the distribution of prediction scores and identifying the ML models that produce abnormal outcomes. Abnormalities were identified by minimal variance, such as *distance-weighted discrimination with Radial Basis Function Kernel* (*dwdRadial*^16^), and overly confident predictions that result in Bernoulli-like outcomes, such as *stochastic gradient boosting* (*gbm*^17^) and *multi-layer perceptron* (*mlp*^18^). Additionally, **Figure 3B** visualises the prediction distribution in held-out samples, which can be distinct compared to training predictions. The ensemble was retrained on 80% of randomly sampled training data and used for prediction on the remaining 20%. In this case, the distribution by *distance-weighted discrimination with a polynomial kernel* (*dwdPoly*^16^) is small relative to the other models.

**Figure 3.**
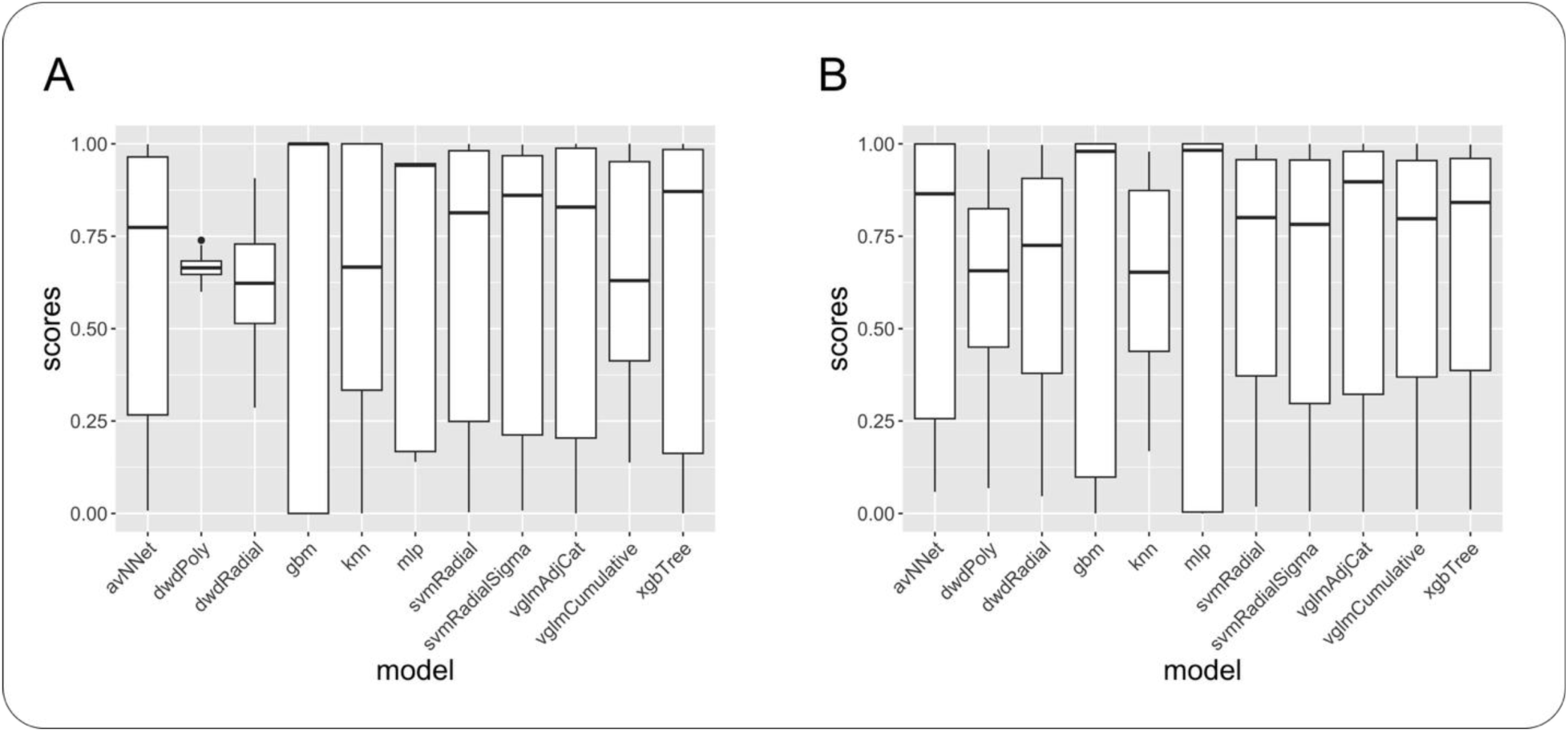
Boxplots of training predictions per model in the ensemble using the optimised panel. The prediction boxplots are presented for (A) in-sample predictions and (B) held-out predictions.

Inconsistencies and unexpected results have been identified in 4 models: *dwdRadial*, *gbm*, *mlp*, and *dwdPoly*. These models have been removed from the ensemble, leaving 7 models: *model-averaged neural network* (*avNNet*^19^), *k-nearest neighbours* (*knn*^20^), *support vector machine with radial basis function kernel (svmRadial* and *svmRadialSigma*)^16^, *adjacent categories probability model for ordinal data* (*vglmAdjCat*^21^), *cumulative probability model for ordinal data* (*vglmCumulative*^21^), and *extreme gradient boosting* (*xgbTree*^22^).

This diagnostic filtering resulted in a final ensemble composed only of models contributing complementary and reliable signals, thereby improving the robustness and interpretability of downstream validation results.

### Internal Validation Using Leave-One-Out Cross-Validation

We assessed the internal performance of the optimised ensemble model using leave-one-out cross-validation (LOOCV) across the European discovery cohorts (EU-cohort 1–3). In this framework, each patient is held out once as an independent test sample while the model is trained on all remaining patients using the optimised panel. The trained model then predicts the held-out patient’s class, yielding fully out-of-sample predictions for every individual in the dataset.

The ensemble demonstrated consistently strong discriminatory performance across all analysis groups (**Table 8**). Sensitivity ranged from 83% to 96%, specificity from 72% to 88%, and AUC values were uniformly high (≥0.94 for EU-cohort 1-2). When all European cohorts were combined, the model achieved 90% sensitivity, 80% specificity, and a mean AUC of 0.94. As expected, these metrics likely represent an optimistic upper bound, as internal validation does not account for cohort- or site-specific shifts encountered during external testing.

**Table 8.** Internal validation (LOOCV) metrics of the ensemble model trained on EU-cohort 1-3 using the optimised panel, stratified by the analysis group/cohort. N/A’s are present in EU-cohort 3 due to the lack of controls. NA, not applicable.

| Cohorts | Accuracy | Sensitivity | Specificity | F1-score | AUC |
| --- | --- | --- | --- | --- | --- |
| EU-cohort 1 | 0.86 | 0.83 | 0.88 | 0.80 | 0.96 |
| EU-cohort 2 | 0.84 | 0.96 | 0.72 | 0.86 | 0.94 |
| EU-cohort 3 | 0.89 | 0.89 | NA | NA | NA |
| All | 0.86 | 0.90 | 0.80 | 0.89 | 0.94 |

### Quasi-external Validation on the European Cohorts

To assess the generalisability of the lipid-based predictive model beyond the European discovery cohorts, we conducted both quasi-external and external validation analyses. Quasi-external validation evaluates performance on held-out European cohorts (EU-cohort 1-2) that were not used for model training but still share similarities with the discovery data. However, because the optimised panel was derived from the full discovery set, this evaluation carries an element of information leakage and therefore represents an optimistic estimate. In contrast, external validation tests the fully trained model on an independent dataset (i.e., the Australian AU-cohort), which is geographically and temporally distinct from EU-cohort 1– 3. This provides a potentially more realistic estimate of generalisability.

For quasi-external validation, we trained models on EU-cohort 2–3 and EU-cohort 1 and 3 and used them to predict outcomes in EU-cohort 1 and EU-cohort 2, respectively. As shown in **Table 9**, performance remained broadly consistent with LOOCV results, with high sensitivity and only modest reductions in AUC. In contrast, specificity decreased, particularly in EU-cohort 1, indicating reduced ability to correctly identify controls in held-out European data. This reduction in specificity explains the corresponding drop in accuracy and F1-score for these groups.

**Table 9.** Quasi-external and external validation of the ensemble model using the optimised panel, stratified by the analysis groups.

| Analysis Group | Accuracy | Sensitivity | Specificity | F1-score | AUC |
| --- | --- | --- | --- | --- | --- |
| Before Platt scaling |  |  |  |  |  |
| EU-cohort 1 | 0.84 | 0.85 | 0.83 | 0.78 | 0.96 |
| EU-cohort 2 | 0.74 | 0.97 | 0.52 | 0.79 | 0.92 |
| AU-cohort | 0.47 | 0.98 | 0.34 | 0.42 | 0.81 |
| After Platt scaling |  |  |  |  |  |
| AU-cohort | 0.66 | 0.76 | 0.64 | 0.47 | 0.81 |

### External Validation on the Australian Cohort

External validation was conducted on the AU-cohort to assess the model’s ability to generalise across geographically distinct populations. As initial exploratory analyses revealed substantial batch separation between European and Australian samples (**Figure 1B**), we applied ComBat^23^ harmonisation to reduce site- and batch-related variability. Batch correction was implemented in a way that avoided information leakage: the European discovery cohorts (EU-cohort 1–3) were used as the reference for estimating batch-adjustment parameters, and these parameters were then applied to the AU-cohort without incorporating any information from the validation labels. This approach preserves the integrity of the training–validation split and ensures that the model developed on the discovery data is evaluated on a properly corrected but still independent external cohort. The batch-corrected UMAP embeddings (**Figure 4A**) show improved alignment between AU-cohort and EU-cohort 1–3, indicating effective mitigation of major batch-driven structure.

**Figure 4.**
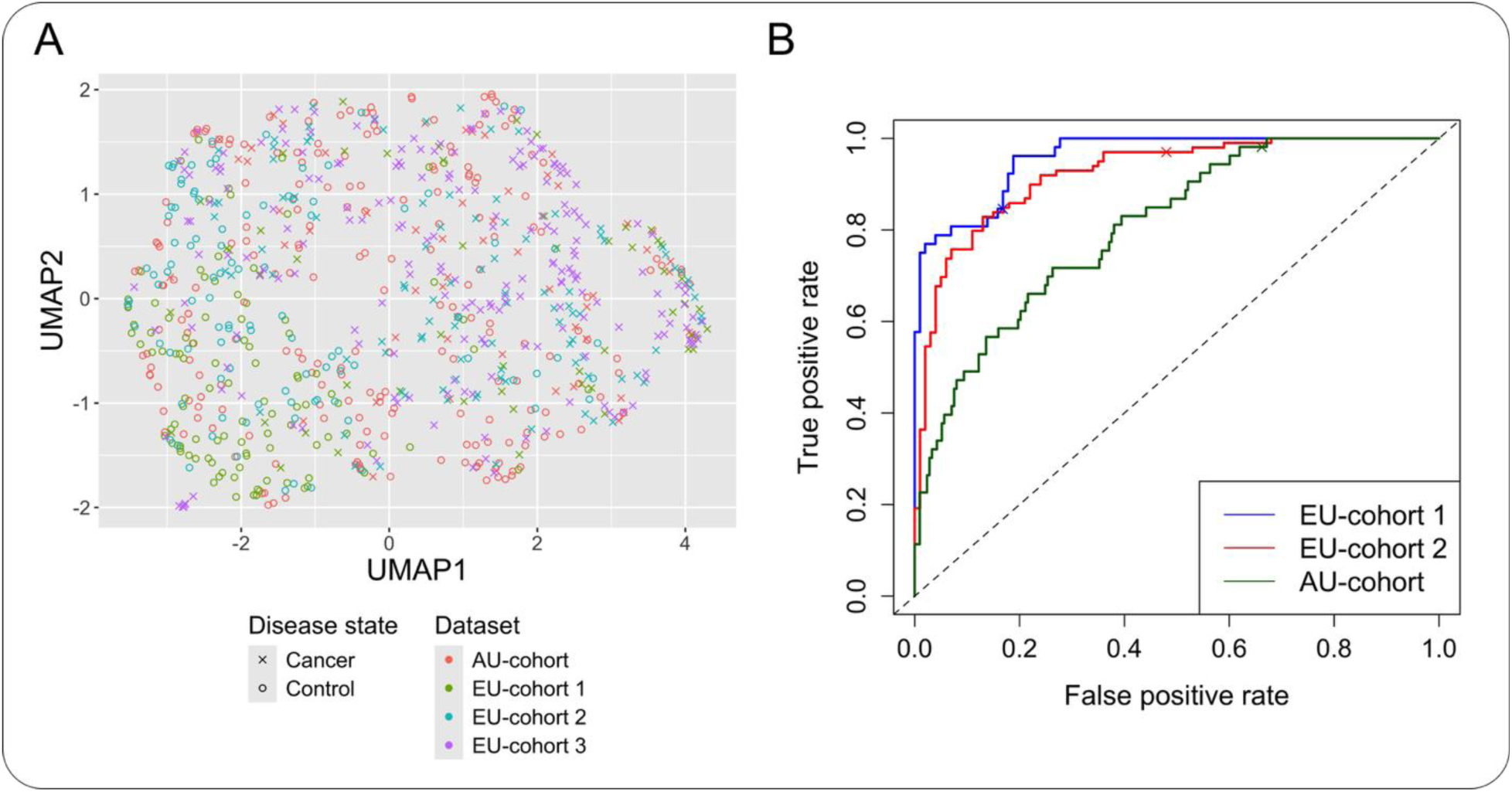
(A) The ComBat batch-corrected UMAP for the indicated cohorts and sample types. (B) Validation cohort ROC curves for the indicated cohorts (EU-cohorts are quasi-external). The cross on each ROC curve indicates the model cutoff of 0.5. ROC, receiver operating characteristic.

The ensemble was then trained on EU-cohort 1–3 and was used to predict patient outcomes in the AU-cohort (**Table 9**). The specificity was lower relative to quasi-external validation, at 34%, while sensitivity remained high at 98%. The model could reasonably discriminate disease states in the Australian cohort, with an AUC reduction of ≤ 15% points (to 81%). The analyses of group-specific ROC curves are shown in **Figure 4B**, highlighting reduced performance of external validation on the AU-cohort compared to the other cohorts. This is likely attributed to information leakage and demographic differences. This imbalance, i.e., high sensitivity paired with low specificity, indicates that the model tends to over-call cancer in the external cohort. As shown in subsequent sections, calibrating the decision threshold can partially correct this imbalance and improve overall diagnostic performance.

### Ensemble Calibration with Platt Scaling

Previous analyses used a model score cutoff of 0.5 to classify samples as case or control and therefore to determine sensitivity and specificity. However, using a fixed decision threshold of 0.5 is unlikely to be appropriate for external validation, particularly given the modest class imbalance in the training data and the marked metric imbalance observed in the Australian cohort prior to calibration (**Table 9**). To obtain a more meaningful balance between sensitivity and specificity, we calibrated the ensemble’s output probabilities using Platt scaling ^24^ and then defined a decision threshold that would achieve approximately 70% sensitivity in out-of-distribution data.

While a simple approach would be to identify the threshold yielding 70% sensitivity on the training predictions, such a strategy does not generalise well and typically amplifies metric imbalance on external datasets. Platt scaling, in contrast, fits a logistic regression model to the ensemble’s out-of-sample *training* predictions, producing calibrated probabilities that are more stable and transferable across populations. The calibrated model was then used to select a threshold aligned with the desired sensitivity target.

After Platt scaling, external validation on the AU-cohort showed substantially improved balance: sensitivity reached 76%, closely matching the target, and specificity increased from 34% to 64%, while the AUC remained stable at 0.81 (**Table 9**). These calibrated metrics reflect a more clinically realistic trade-off between detecting cancers and minimising false positives in an out-of-distribution population. Although performance remains lower than in quasi-external validation, this is expected due to demographic differences and residual domain shift. Given its ability to stabilise probability estimates and rebalance diagnostic metrics, Platt scaling was applied in all downstream analyses.

### Refinement of the optimised Panel

The initial optimised panel, derived using the Boruta feature-selection approach, consisted of 15 lipids. While Boruta identifies features with consistent discriminative value, it operates as a heuristic and does not guarantee that the resulting panel is minimal, free of redundancy, or optimal for downstream assay development. To address these considerations, we generated and evaluated three refined subsets of the optimised panel: a minimal viable panel, a low-collinearity panel, and an analytically viable panel suited for potential translational deployment as a clinical assay with commercial potential (**Table 6**). All panels have been compared to the performance of the optimised panel.

#### Minimal viable panel

To identify the smallest set of lipids capable of maintaining acceptable performance, we applied recursive feature elimination (RFE)^25^ with leave-group-out validation. Lipids were ranked according to their contribution to model performance, and the ensemble’s 5-fold cross-validated AUC was recorded as lipids were progressively removed (**Figure 5A**). The AUC remained stable after exclusion of the first 10 lipids but dropped sharply when PC 18:1/20:4 was removed, defining a minimal viable subset of six lipids (**Table 6**). Bootstrapped external validation on the AU-cohort (**Table 10**) showed that this reduced panel maintained comparable sensitivity to the optimised panel (72%, +2%), but with expected trade-offs in specificity (–5% points) and AUC (–3% points).

**Figure 5.**
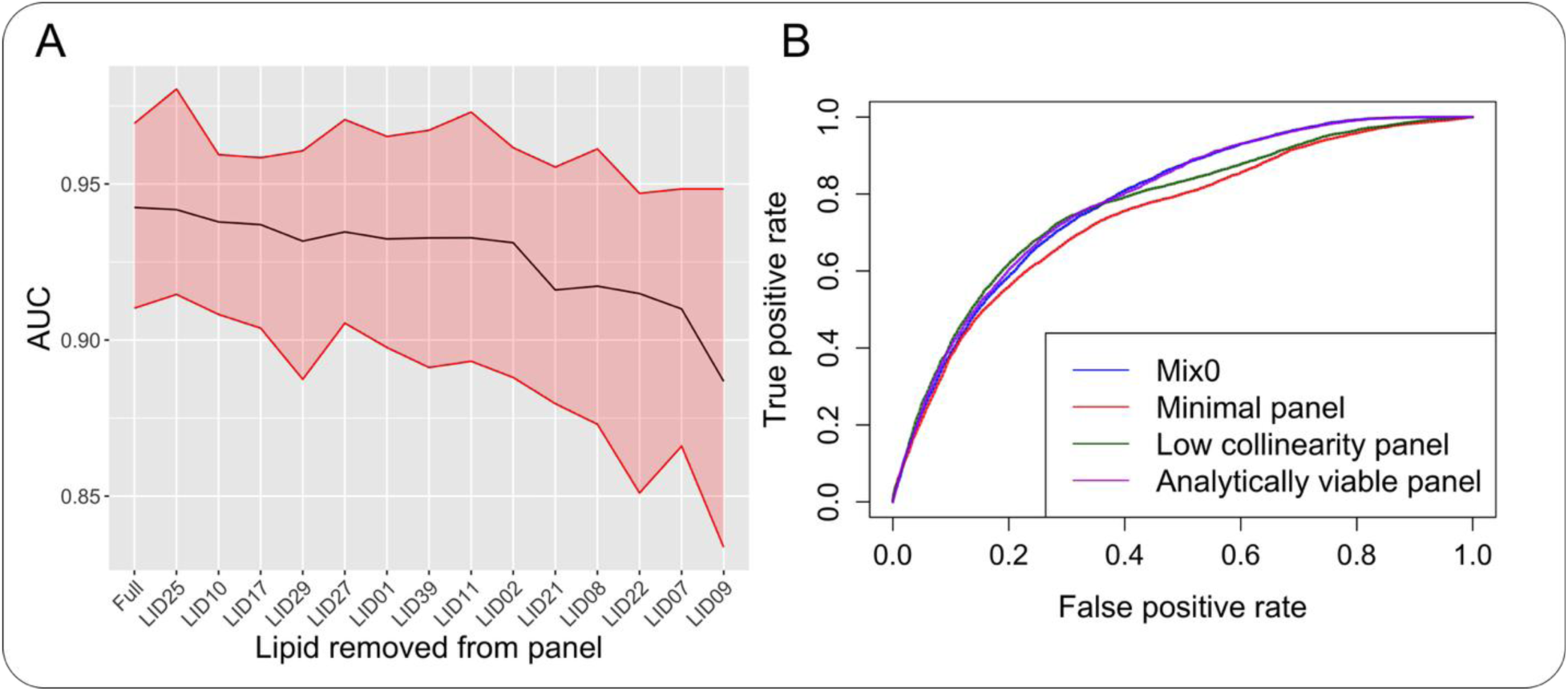
(A) The evolution plot demonstrates the decrease in AUC as lipids are progressively removed in the order given by recursive feature elimination. (B) Set of ROC curves comparing the predictive performance of the optimised panel and its candidate subsets. ROC, receiver operating characteristic.

**Table 10.** Bootstrapped external validation metrics for each panel. A Platt-scaled ensemble was trained on 100 bootstraps of the training dataset and tested on the AU-cohort. The median of each metric is presented alongside the 95% bootstrap confidence interval in parenthesis.

| Panel | Accuracy | Sensitivity | Specificity | F1-score | AUC |
| --- | --- | --- | --- | --- | --- |
| Optimised | 0.72<br>(0.67 - 0.77) | 0.70<br>(0.60 - 0.81) | 0.72<br>(0.65 - 0.81) | 0.50<br>(0.46 - 0.54) | 0.79<br>(0.77 - 0.82) |
| Minimal viable | 0.68<br>(0.62 - 0.74) | 0.72<br>(0.60 - 0.77) | 0.67<br>(0.59 - 0.76) | 0.46<br>(0.43 - 0.50) | 0.76<br>(0.74 - 0.78) |
| Low collinearity | 0.70<br>(0.64 - 0.75) | 0.76<br>(0.65 - 0.81) | 0.69<br>(0.61 - 0.76) | 0.50<br>(0.45 - 0.54) | 0.78<br>(0.75 - 0.80) |
| Analytically viable | 0.72<br>(0.66 - 0.76) | 0.70<br>(0.62 - 0.81) | 0.72<br>(0.64 - 0.78) | 0.50<br>(0.46 - 0.55) | 0.79<br>(0.76 - 0.82) |

#### Low-collinearity panel

As correlated features have the potential to introduce redundancy and inflate model variance, we constructed a low-collinearity subset by removing lipids with pairwise correlations ≥0.8 (**Figure 2D**). This procedure eliminated LPC 16:0 and LPC 14:0, yielding a 13-lipid panel (**Table 6**). Interestingly, external validation showed improved sensitivity (76%, +6%) with only a modest reduction in specificity (69%, –3%) (**Table 10**). The overall AUC (0.78, −1%) remained comparable to the optimised panel, demonstrating that reducing feature redundancy can enhance certain aspects of model performance.

#### Analytically viable panel

For a lipid assay to be clinically deployable, analytical parameters are critical. Therefore, we removed lipids with lower inter-assay precision, linearity limitations, or confounding isobaric interference, specifically, PC 18:1/20:4. The resulting analytically viable panel achieved similar performance on the AU-cohort (**Table 10**).

#### Comparison of refined panels

Although performance differences between panels are expected, their confidence intervals largely overlap, illustrating a degree of uncertainty and indicating that no subset is statistically inferior across all metrics. The ROC curves (**Figure 5B**) further elucidate the similarity in each panel. However, the *analytically viable panel* meets essential assay development requirements, making it the most suitable candidate for further translational development.

### Assessment of Predictions in Relation to Clinical and Patient-Level Variables

The primary performance metrics reported thus far are based solely on lipid-derived predictions of cancer status. However, lipid profiles can be influenced by patient-level clinical characteristics. For a biomarker-based diagnostic to be clinically useful, its predictions should ideally be independent of factors such as age, tumour characteristics, and metabolic status, otherwise, confounding effects may undermine its generalisability. To evaluate this, we examined whether the ensemble prediction scores were associated with key clinical features in the external AU-cohort.

We stratified AU-cohort by cancer status and fitted linear models with ensemble prediction scores as the response and each clinical feature as the predictor. Analysis of variance (ANOVA) was then conducted to determine whether the ensemble predictions are significantly different across the different values of the clinical feature (**Figure 6)**.

**Figure 6.**
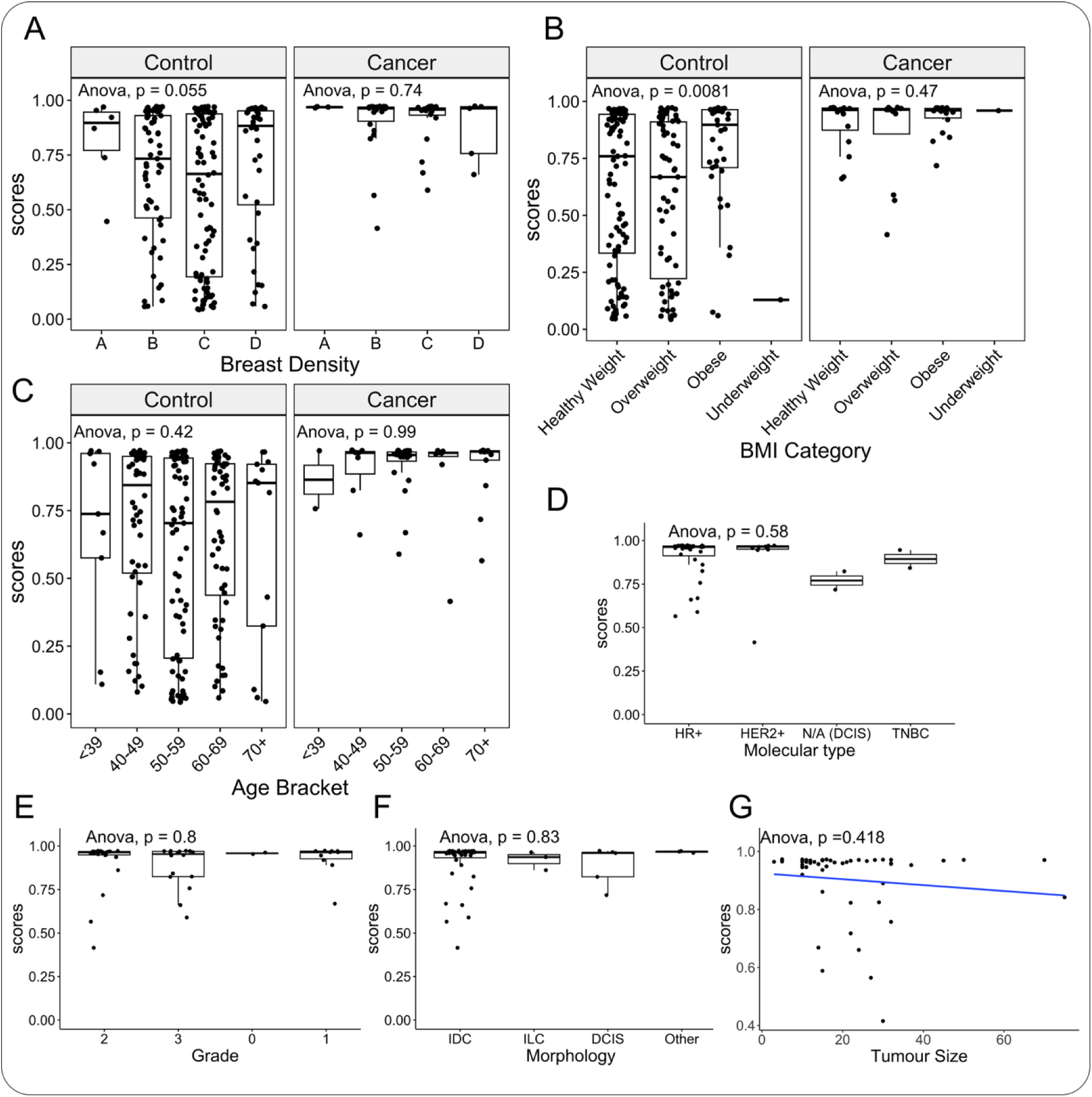
The distribution of prediction scores across important clinical features, including (A) breast density, (B) BMI category, (C) age bracket, (D) molecular type, (E) cancer grade, (F) cancer morphology, and (G) tumour size.

Breast density is a well-known confounder for breast mammography, as diagnostic sensitivity is reduced in women with dense breasts ^7,8^. The ANOVA indicates that there is no statistically significant difference in the mean prediction across different breast densities (**Figure 6A**). This suggests that the lipid-based panel captures biological signals independent of breast density, offering a potential advantage over imaging-based screening.

Age and body mass index (BMI) are physiological factors known to influence lipid metabolism. Although our analysis indicates minimal differences across age brackets, the prediction means differ across BMI categories in control patients, likely due to changes in lipid concentrations attributed to lifestyle, diet, and metabolic shifts (**Figure 6B and 6C**). Specifically, control patients classified as obese exhibited higher prediction scores, making them more likely to be misclassified as cancer. This is reflected in performance metrics: obese patients had statistically significantly lower specificity (60%) compared with overweight individuals (82%) and the bootstrap confidence intervals tended to be lower in obese patients than in healthy-weight patients, as detailed in **Table 11**. Sensitivity, however, remained stable across BMI categories, indicating that the reduction in performance is largely driven by misclassification of obese controls.

**Table 11.** The bootstrapped external validation metrics for the analytically viable panel, stratified by BMI categories. The median and 95% bootstrapped confidence intervals are presented for each metric in parentheses.

| BMI | Accuracy | Sensitivity | Specificity | F1-score | AUC |
| --- | --- | --- | --- | --- | --- |
| Healthy weight | 0.70<br>(0.64 - 0.76) | 0.69<br>(0.56 - 0.75) | 0.71<br>(0.64 - 0.79) | 0.39<br>(0.33 - 0.44) | 0.79<br>(0.75 - 0.82) |
| Overweight | 0.81<br>(0.74 - 0.87) | 0.75<br>(0.67 - 0.75) | 0.82<br>(0.74 - 0.89) | 0.55<br>(0.47 - 0.64) | 0.80<br>(0.76 - 0.83) |
| Obese | 0.64<br>(0.57 - 0.67) | 0.72<br>(0.61 - 0.89) | 0.60<br>(0.50 - 0.68) | 0.57<br>(0.49 - 0.64) | 0.73<br>(0.69 - 0.78) |

Additionally, among patients with confirmed breast cancer, the model’s prediction scores were not significantly associated with molecular subtype, tumour grade, morphology, or tumour size (**Figures 6D–6G**). This indicates that the assay detects cancer-associated lipid perturbations largely independently of disease subtype or stage-related characteristics, a desirable property for an early detection test.

Overall, these findings suggest that while the lipid panel is broadly robust to many clinical and tumour-related variables, metabolic factors associated with higher BMI may shift lipid profiles in ways that resemble the cancer phenotype. The observed reduction in specificity in obese control patients, therefore, appears to be driven by shifts in the underlying metabolic landscape rather than tumour-related effects. This observation highlights BMI as a potential confounding factor in lipid-based modelling and motivates future methodological work to more explicitly account for metabolic heterogeneity when developing and refining predictive models. Further expansion of the training dataset to include more metabolically diverse populations may also enhance performance in this subgroup.

### Confidence-Stratified Performance of the Predictive Model

The performance metrics, such as sensitivity, specificity, accuracy, F1-score, and AUC, summarise how well a model performs across an entire population. However, they do not convey how *confident* the model is in any given prediction. Because the ensemble was calibrated using Platt scaling, its output probabilities approximate calibrated likelihoods and can therefore be interpreted as an estimate of prediction confidence ^26^. In a clinical setting, this confidence information may have practical value: low-confidence classifications may prompt further diagnostic workup, whereas high-confidence predictions could strengthen clinician trust in an assay. Alternatively, if assay performance is greatly enhanced with confident predictions, it could support ongoing research that focuses on understanding why low-confidence predictions caused poorer assay performance. This, in turn, could provide an opportunity to use additional diagnostic algorithms for low-confidence predictions or further research into why such samples were predicted with low confidence.

To explore this dimension, we evaluated the *analytically viable panel* under a confidence-stratified framework (**Table 12**). Predictions were grouped into 1) Confident predictions, defined as moderate to high confidence (scores ≥0.33), and 2) High-confidence predictions (scores ≥0.66). The composition of prediction confidence and the ROC curve for each group is presented in **Figure 7**.

**Figure 7.**
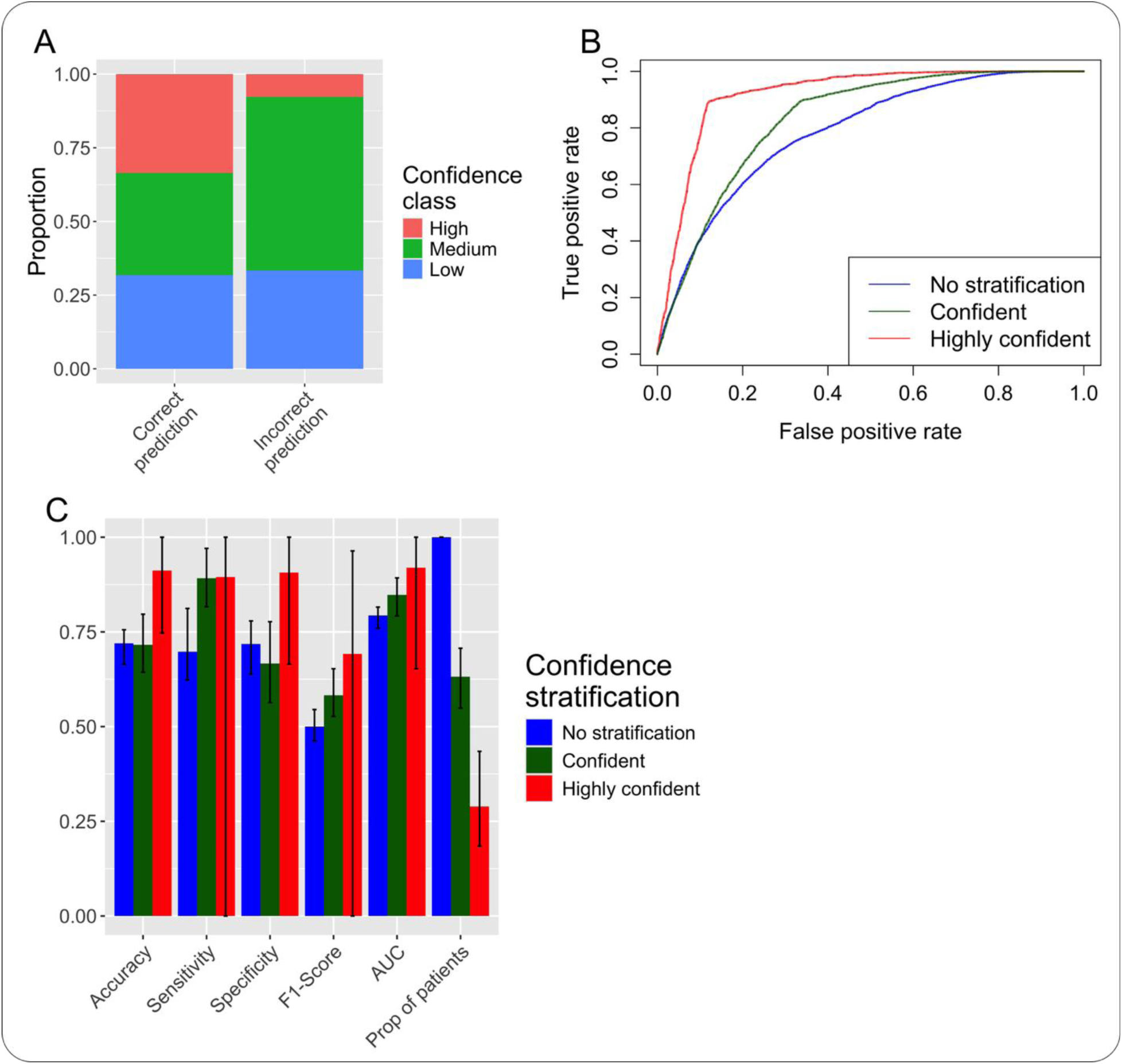
(A) Certainty level of predictions, stratified by correct and incorrect predictions. (B) ROC curves for all predictions split into the indicated confidence categories. No stratification, all samples. (C) Median and 95% bootstrap confidence intervals for each indicated metric.

**Table 12.** Bootstrapped external validation metrics for the analytically viable panel, stratified by confidence bands. The median and 95% bootstrapped confidence intervals are presented for each metric in parentheses.

| Stratification | Accuracy | Sensitivity | Specificity | F1-Score | AUC | Proportion of patients |
| --- | --- | --- | --- | --- | --- | --- |
| **Confident**<br>**($\geq 0.33$)** | 0.72<br>(0.64 -<br>0.80) | 0.89<br>(0.82 -<br>0.97) | 0.67<br>(0.56 -<br>0.78) | 0.58<br>(0.53 -<br>0.65) | 0.85<br>(0.79 -<br>0.89) | 0.63<br>(0.55 -<br>0.71) |
| **Highly**<br>**confident**<br>**($\geq 0.66$)** | 0.91<br>(0.75 -<br>1.00) | 0.90<br>(0.00 -<br>1.00) | 0.91<br>(0.66 -<br>1.00) | 0.69<br>(0.00 -<br>0.89) | 0.92<br>(0.65 -<br>1.00) | 0.29<br>(0.18 -<br>0.44) |

### Moderate-to-high confidence predictions

Approximately 63% of patients fell within the moderate-to-high confidence category. Within this subset, median sensitivity, F1-score, and AUC increased by 19%, 8%, and 6% points, respectively, compared with the full test population. Specificity decreased modestly (−5%), and accuracy remained unchanged. Importantly, the confidence intervals for sensitivity were significantly different from those of the full cohort, indicating that the model is particularly reliable when assigning a confident cancer prediction. The 97.5% quantile of the AUC is significantly higher than that of the full cohort AUC, whereas the corresponding 2.5% quantiles are nearly equal. Together, these observations indicate that the model performs equivalently or better when making confident predictions (**Table 12**).

### High-confidence predictions

A smaller proportion of samples (29%) met the high-confidence threshold. In this subset, median performance improved further—sensitivity (+20%), specificity (+19%), and AUC (+13%). This gave sensitivity of 90%, specificity of 91% and an AUC of 92% (**Table 12**). However, the wide confidence intervals reflect the reduced sample size, limiting the statistical certainty of these improvements. Clinically, this indicates that while the strongest predictions are highly reliable, the number of patients reaching this confidence threshold may be too small to guide routine decision-making without additional research.

Overall, confidence stratification provides insight into how the model behaves when it is certain versus uncertain, aligning with the concept of “selective prediction” or “abstention” in medical AI. The observed performance differences across confidence strata highlight that uncertainty-aware outputs can be informative for understanding model limitations and strengths. Notably, a substantial proportion of samples (37%) fall below the predefined confidence threshold, underscoring current constraints related to data scale, cohort diversity, and model calibration. These findings help identify clear directions for future work, including dataset expansion, improved uncertainty modelling, and calibration strategies, with the aim of increasing the robustness and consistency of high-confidence predictions in subsequent model iterations.

## Discussion

In this work, we developed and validated a plasma lipid–based, machine-learning-enabled assay for early breast cancer detection across multiple, geographically distinct cohorts. Starting from 48 lipids with identify confirmation and absolute quantitation, we applied a robust, data-driven feature-selection pipeline to derive a 15-lipid optimised panel capable of distinguishing cancer from control samples with high accuracy in European cohorts. Internal LOOCV and quasi-external validation demonstrated consistently strong diagnostic performance, with AUC values exceeding 92% across all discovery datasets.

External validation using the Australian AU-cohort revealed expected reductions in performance due to pronounced batch and demographic differences. By integrating ComBat batch harmonisation and Platt-scaled calibration, we achieved a clinically meaningful balance of 76% sensitivity and 64% specificity, with an AUC of 81%. These results demonstrate that although cross-population generalisation remains challenging, lipid-based signatures retain substantial diagnostic signal across sites.

To support translational feasibility, we explored refinement of the optimised panel through recursive feature elimination, collinearity assessment, and analytical suitability. Among the resulting subsets, the analytically viable panel offered the most favourable combination of diagnostic performance, assay robustness, and therefore a potential to develop the assay towards a clinical assay at scale.

Importantly, predictions were largely independent of key tumour characteristics and patient factors, including breast density, tumour size, molecular subtype, grade, and morphology, highlighting the biological specificity of the lipid signature. BMI emerged as a notable exception, with obese control patients exhibiting higher prediction scores, likely reflecting metabolic alterations that partially resemble the cancer-associated lipid profile. This finding underscores the need for additional adjustments or confirmatory testing in metabolically heterogeneous populations. Confidence-stratified analyses further revealed that the model performs particularly well when predictions are made with high confidence, yielding up to 90% sensitivity, 91% specificity and an AUC of 0.92. While only a subset of patients currently falls within this high-confidence range, these findings suggest that calibrated probability outputs may be leveraged to guide reflex testing or risk-stratified clinical pathways. Moreover, these findings point towards a focus area for ongoing research to improve the confidence of model predictions.

Collectively, our results demonstrate that plasma lipidomics, integrated with calibrated machine-learning models, provides a promising avenue for non-invasive breast cancer detection. Incorporating more diverse training data—particularly from Australian and metabolically varied populations—will be essential for further reducing domain shift, improving specificity, and consolidating the biomarker panel. With expanded datasets and continued assay refinement, this approach has potential to support the development of scalable, blood-based early detection strategies that complement existing imaging-based screening pathways.

## Materials and Methods

### Blood samples

Fasted blood samples were prospectively collected throughout 2020–2023 from female participants at multiple sites in Eastern Europe and Australia. The presence of breast cancers in treatment naïve patients was confirmed by tissue biopsy. Healthy controls had not been previously diagnosed with breast cancer or any other cancer, except successfully treated non-melanoma skin cancers. The collected blood samples were kept at 4-8 °C and processed into plasma. Samples were stored at −80 °C until analysis. Cohorts included healthy controls and individuals with invasive ductal carcinoma (IDC), invasive lobular carcinoma (ILC) and ductal carcinoma in situ (DCIS). Breast cancers were also stratified by molecular subtype based on surface receptor staining, as HR+ (hormone receptor positive; estrogen or progesterone receptor, ER/PR), HER2+ (HER2 positive) or TNBC (Triple Negative Breast Cancer; ER, PR and HER2 negative). QC plasma was obtained from the Australian Red Cross LifeBlood (Alexandria, NSW, Australia). This study was conducted in accordance with the Declaration of Helsinki. Recruitment of subjects in Europe was approved by the Institutional Review Board of Ochsner Health, New Orleans, LA, USA (protocol code 2015.101). The sponsor of the biospecimen collection study was Capital Biosciences, Gaithersburg, MD, USA. Recruitment of subjects in Australia was approved by the Sydney Local Health District Human Research Ethics Committee (protocol code 2020/ETH02051).

### LC-MS/MS materials and methods

The methods used to conduct the plasma lipid measurements have been described^14^. Briefly, a two-phase lipid extraction method was used to isolate lipids from plasma samples. A targeted LC-MRM method was performed using a SCIEX Exion UHPLC system coupled to a 7500 triple quadrupole mass spectrometer (SCIEX, Ontario, Canada). Reference standards and stable isotope-labelled internal standards were used to accurately quantify target lipids (ANTO, Avanti Polar Lipids, Cayman Chemical, Echelon Biosciences, Sigma Aldric, TRC and Precion Inc.).

### Data pre-processing

The data consists of four Cohorts, the EU (EU-Cohort 1-3) and AU (AU-Cohort) Cohorts. The datasets were log10-transformed, and missing data were minimum imputed using the EU Cohort.

### Feature selection protocol

Feature selection was conducted using Boruta. It trains a random forest on both the original features and noisy variants, referred to as shadow features. A feature was deemed important if it could separate cancer and control states better than its shadow variant.

Boruta is a heuristic; it has multiple hyperparameters that can impact the final selection. The number of trees used in Boruta was set to 500, which controls the variance of the features that have been selected. The maximum number of importance source runs was set to 2500, which increases the confidence of the selected features by allowing more iterations to resolve tentative selections. The p-value threshold has been set to the default value of 0.01.

Boruta was repeated over 1000 bootstraps to identify lipids that are consistently selected, thereby improving the panel’s robustness. Lipids that were selected in more than 70% of bootstraps were chosen as the optimized panel.

### Establishment of the ensemble

An ensemble of models constitutes multiple models, enhancing prediction outputs by model averaging. The performance improvement is dependent on model complementarity: the model’s learning patterns must be sufficiently different to provide additional insights into the data. Additionally, models that output prediction probabilities have been chosen to assess the confidence of each prediction. The models are as follows:

#### Support Vector Machine with Radial Basis Function Kernel

A statistical learning method that uses margin-based approaches to separate data. It leverages support vectors – data points that determine the separating hyperplane – by calculating a maximal margin that separates them. Nonlinearity is introduced with the kernel function.

#### Distance-weighted discrimination with Radial Basis Function Kernel and distance-weighted discrimination with Polynomial Kernel

Like SVM, a separating hyperplane is calculated; however, the margin is calculated by minimizing the sum of inverse distances to every data point.

#### Boosting models

Models that iteratively combine base models, such as decision stumps, so that each iteration improves the predictions of the past models. *Gbm* and *xgbTree* are examples of such algorithms. *Gbm* utilizes a bagging procedure by inducing randomness in the learning task to improve model performance. *XgbTree* is a highly scalable boosting algorithm that incorporates advanced regularization techniques and second-order approximations of the gradient to improve accuracy.

#### Vector Generalized Linear-based models

General statistical models that fit a linear model to the data.

#### Neural Networks

Models consisting of neuron layers that recognize underlying patterns in the data, enabling the learning of complex functions. In this study, simple feedforward networks, such as *mlp*, are preferred to reduce overfitting. *AvNNet* is an alternative network that leverages bagging and model averaging to reduce overfitting.

#### K-nearest neighbours

A heuristic method that assigns classes to data points based on the class of their neighbours. The performance is dependent on distance metrics and the cardinality of neighbours.

### Training and refinement of the ensemble

The individual models were trained on EU-cohort 1-3 with the assistance of hyperparameter optimization in *caret’s train* function^27^. Specifically, the optimization method is administered by the *trainControl* function, which has been configured to generate a random hyperparameter space. Each hyperparameter was tested in 10 iterations of leave-group-out cross-validation (LGOCV) – trained and tested on 80% and 20% of the data, respectively. The hyperparameter that conferred the highest LGOCV AUC was used to train the final model. The patient outcome was produced by averaging the model predictions.

Although the models have been carefully selected based on their attributes, they may not perform well in practice. Models that produce minimal prediction variance are problematic. Therefore, each model was investigated by plotting a boxplot of their training predictions. Furthermore, boxplots for out-of-sample predictions have also been generated, as their predictive distributions can differ from in-sample distributions.

### Evaluation design and metric reporting

The selected lipids will be validated with the optimized ensemble under different test schemas.

#### Leave-one-out cross-validation

An internal validation approach involving the training set, which provides an optimistic estimate of test metrics on unseen data. The ensemble was trained on all but one sample, which was used as the test. This was repeated for each sample, generating out-of-sample predictions per patient, which were used to calculate *leave-one-out cross-validation* metrics.

#### External validation

An approach that validates a trained model on unseen data and tests the generalizability of the model. The unseen data is ideally temporally and geographically distinct. The trained model was used to predict the disease state of unseen data, and the external validation metrics were calculated.

#### Bootstrapped external validation

An approach that, like the non-bootstrapped version, highlights the generalizability of the model. However, multiple models were trained on bootstraps of the training data. The models were tested on unseen data, which was used to generate distributions on each test metric. The median value and 95% bootstrapped confidence intervals can be reported to make statistical assertions regarding performance improvements.

### Calibrating the optimized ensemble by Platt scaling

The optimized ensemble consists of 7 models, which are poorly calibrated. This means that the prediction probabilities do not reflect the chance of observing the disease state in our datasets. This can cause metric imbalance. *Platt scaling* calibrates the ensemble, which results in a balanced metric readout.

In this study, Platt scaling has been applied to calibrate out-of-sample training predictions, enabling the calculation of a threshold that achieves 70% sensitivity in unseen data. The threshold was set at 70% sensitivity to ensure a balance in the specificity. The training data was randomly split into 5 folds; 4 folds were used for training the ensemble, which was used to predict on the remaining held-out fold. The held-out predictions were calibrated with logistic regression. This process was repeated for each fold, resulting in two lists of held-out predictions before (1) and after calibration (2). A logistic regression was trained on (1). This model will be used to calibrate ensemble predictions on unseen data. Additionally, the 70% sensitivity threshold was calculated using (2). Thus, for incoming data, the ensemble will be used for prediction, followed by calibration using the logistic regression trained by (1). The threshold will then be used to determine the class.

### Generation of the minimal viable panel with recursive feature elimination

The minimal viable panel was determined by using RFE. RFE is a heuristic that performs backwards selection on a feature set to remove features sequentially in order of performance impact. At each iteration, leave-group-out cross-validation metrics were generated on 50 samples from the training dataset, which had been split so that each sample consisted of 80% training and 20% validation. The worst-performing feature – the one that reduced the performance the most – was removed in the next iteration. RFE returns a list of lipids sorted by their contribution to model performance. An *AUC evolution* plot can then be generated by plotting the 5-fold cross-validated performance as lipids are removed in the order provided by RFE.

### Confidence stratification

The reported metrics can be interpreted as the average performance over the entire test dataset. However, it does not consider the performance of confident predictions, which, in isolation, are predicted well.

Prediction confidence is determined by calculating the ratio of the distance between the prediction and the threshold, and the distance between the threshold and 1 or 0 (if the prediction is above or below the threshold, respectively). Or equivalently,

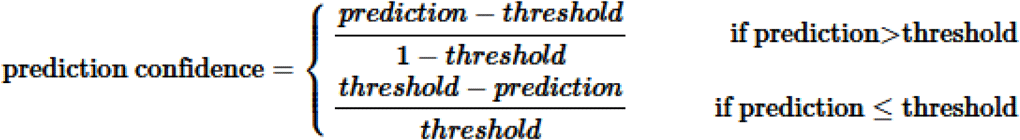

Prediction confidence was then stratified into three categories: Low confidence, corresponding to values lower than 0.33; medium confidence, corresponding to values between 0.33 (inclusive) and 0.66; and finally, high confidence, corresponding to values greater than or equal to 0.66. Equivalently,

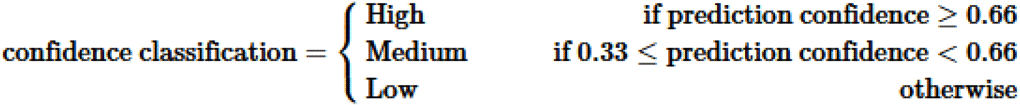

## Data Availability

All data produced in the present study are available upon reasonable request to the authors

## Notes

### Competing Interest Statement

The authors have declared no competing interest.

### Funding Statement

This study was funded by BCAL Diagnostics Ltd, Sydney NSW 2113, Australia

### Author Declarations

This study was conducted in accordance with the Declaration of Helsinki. Recruitment of subjects in Europe was approved by the Institutional Review Board of Ochsner Health, New Orleans, LA, USA (protocol code 2015.101). The sponsor of the biospecimen collection study was Capital Biosciences, Gaithersburg, MD, USA. Recruitment of subjects in Australia was approved by the Sydney Local Health District Human Research Ethics Committee (protocol code 2020/ETH02051).

